# Dynamical Pool-Size Optimization for the SARS-CoV-2 PCR Test

**DOI:** 10.1101/2021.01.12.21249581

**Authors:** Juri Smirnov, Thomas Fenner

## Abstract

In this work, we put forward a novel test strategy, that allows to significantly increase the test capacity for SARS-CoV-2. The test strategy is based on an a priory risk assessment scheme, that allows to dynamically find and adapt an optimal clustering size of test pools. We, furthermore, suggest a method to overcome the efficiency loss of test clustering by avoiding concentration losses in the test samples. We validated our method with several thousand probe pools performing RT-PCR tests, and found it highly effective.

## Introduction

At present, there is an urgent need for an increase in the testing capacity of the molecular biological testing of oropharyngeal swabs for the new SARS-CoV-2 [1]. The effort is hindered by a limited capacity of chemical agents, instruments, and medical personnel to test symptomatic patients. Also, there is a growing need for the testing of personnel in critical professions, such as doctors, firefighters, and the police force, but also business travelers. After being exposed to SARS-CoV-2 positive individuals they need to be quarantined or tested. The second option, assuming a negative test result, allows a quick return to active duty. Thus an increase of test capacities by pooling of patient swab tests seems to be a viable strategy. Pooling techniques are currently used to ensure antiviral quality controls of donated blood samples, see for example Ref. [2]. These pooling methods are effective, since the types of molecular biological tests used are highly sensitive, even to minuscule amounts of viral material in the sample [3].

Different strategies are possible in principle that are based on test pooling [4, 5], however, under realistic laboratory conditions, the logistical effort for the handling of probes has to be minimal to avoid delays and errors in a high throughput environment. We have thus implemented a simple and effective method detailed below in our study. In our initial test, a certain number of swabs from different patients is unified in a single pool. In case the test is negative, all patients from the pool can be considered as negatively tested. In the case the pool test gives a positive result, a second test is needed in which all patients are tested individually. Thus the pooling process has to take place in the diagnostic laboratory, as a second probe sample has to be kept for each patient.

The challenges that arise by pooling test samples of SARS-CoV-2 patients have been discussed in Ref. [6]. In our work, we develop a comprehensive adaptive strategy for pool size optimization and address the issue of sensitivity loss due to dilution processes. This provides a concrete actionable proposal for increasing the number of SARS-CoV-2 tests, especially during the currently ongoing SARS-CoV-2 pandemic.

To conserve cost and resources by a test pooling method the number of patients in a test pool has to be chosen such, that all positive patients are detected, but at the same time the number of test repetitions is kept to a minimum. The distribution of SARS-CoV-2 positive patients is currently inhomogeneous. During the progression of the SARS-CoV-2 pandemic, the fraction of infected individuals can evolve differently in different regions. Also among individuals living in institutions, such as nursing homes, or medical personnel, the numbers of infected individuals can vary. However, well-defined cohorts are likely to have specific mean probability values for the number of infected individuals. For each hypothesis of the underlying average number of infections, an optimal pool size can be defined.

In this work we consider several patient cohorts, identify their specific infection risk factors, and propose optimal pool sizes. Since our method relies on cohort specific risk factors, that rescale the global risk of infection to specific risk within a cohort, our results can be used for risk estimates and pool size adaptation given an evolving total infection probability during the ongoing pandemic. In addition to the theoretical proposal, we perform an experimental validation of the method among a well-defined test cohort.

In Fig. 1, we show a simple example of a cohort test with an underlying positive rate of 3% and demonstrate how an optimal pool size choice outperforms an inefficient pool size choice. In this letter, we present a pool size optimization that can lead to a significant reduction of tests per patient and can thus allow the screening of much larger patient numbers. We note that a test pooling should be only performed if it leads to a conservation of resources and always be communicated and coordinated with the local health agency. We describe the particular pooling method in the following.

**FIG. 1.**
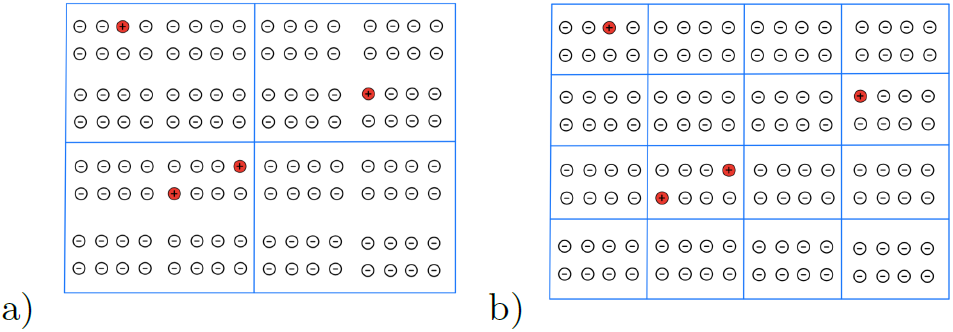
Two different pool size choices for a pool test. The chosen cohort has a 3% infection rate. In panel a) 32 patients are cleared with an initial pool test and 96 tests need to be repeated. In total 100 tests are used to test 128 patients. In panel b) 104 patients are cleared with 16 initial pool tests and 24 tests need to be repeated. In total 40 tests are used to test 128 people. The pool size of 8 in panel b), given the 3% infection rate, is optimal.

The PCR test for SARS-CoV-2 detection is extremely sensitive, even to very low amounts of viral material [7, 8]. We make use of the inherent low false-negative rate in the following. Thus if a given pool test leads to a negative result in the PCR test, all patients from that pool can be considered as negative. In the case that the test is positive, the PCR test is repeated for all patients in that pool individually. From the logistics point of view, this means that the probe material of each patient has to be divided into two upon reception. Overall we will show that in particular in the case that the positive rates are relatively low a significant increase in test efficiency can be achieved. We will present a data set, collected over the past seven months, and demonstrate that we were able to achieve a 79% increase in efficiency in the tested patient cohort.

## Sensitivity and Specificity of Pool Testing

An important question to be addressed is how to test sample unification affects the sensitivity and specificity of a given method. In particular, since the viral load experiences a large day-to-day variation it is important for a testing method to be sensitive to low viral loads, in order not to generate an unacceptable number of falsenegative results.

The primer design of the PCR SARS-CoV-2 test [8] is highly specific. Thus the false positive rate of the method is essentially zero and obviously can not be affected by the unification of several test samples in a pool. The only way a false positive result can be generated is probe contamination during the preparation process, which is a systematic and not stochastic error of the method [9]. On the other hand, the question of sensitivity is much more subtle.

Figure 2 shows the time series of viral loads in pharyngeal swab samples determined by PCR amplification [10, 11]. The significant observation is the large variability of the CT-values in the active phase of Covid-19 infection. The average one-standard deviation interval of CT-values, i.e. the range in which the CT-value falls with a 68% probability spans across 9 Ct-values steps. This corresponds to an average fluctuation of the viral load by a factor of 500. About 10% of the samples from patients in the active phase of Covid-19 are below the detection threshold. This value is an estimate of the underlying systematic uncertainty, most probably connected to the quality of sample collection.

**FIG. 2.**
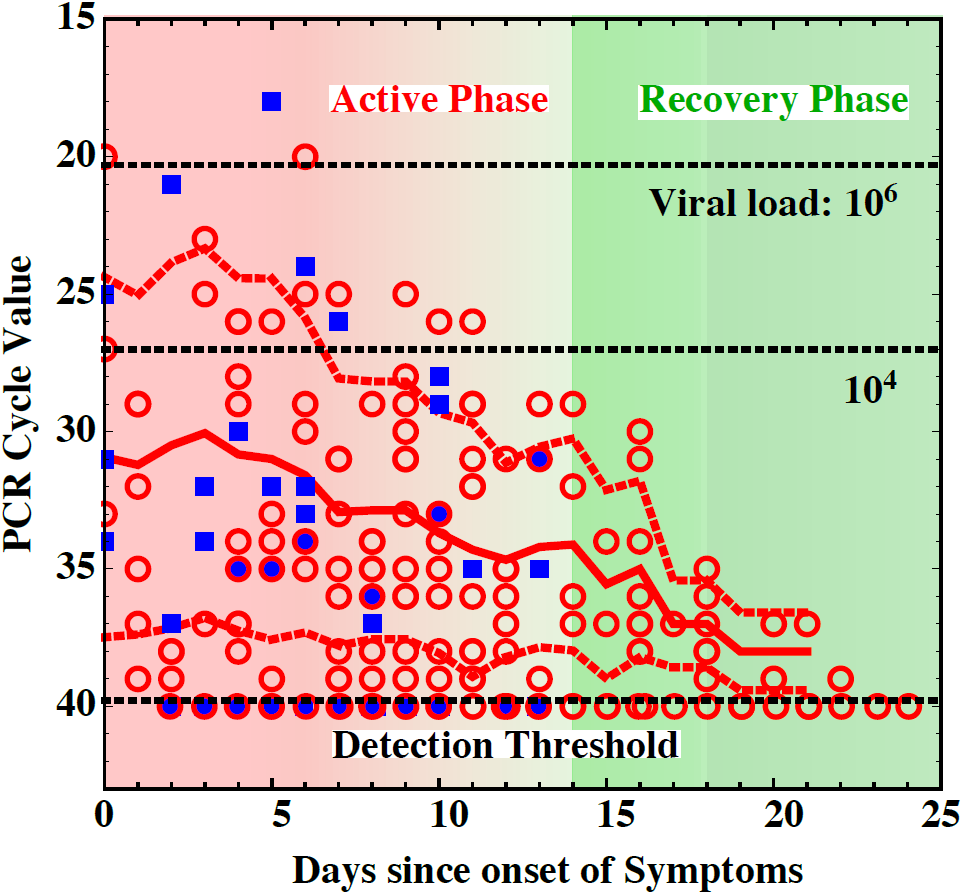
The viral load of pharyngeal swabs, determined by PCR amplification. Panel a) shows a times series of 104 patients from the Wuhan region [10, 11] All tested positive with the SARS-CoV-2 and all were symptomatic. The viral load is logarithmically linked to the number of PCR cycles, after which the probes were detected as positive. The red solid line shows the time evolution of the mean CT-value, and the red dashed lines indicate the one standard deviation contours.

Finally, it becomes obvious from Fig. 2 that given that a patient is in a late stage of the Covid-19 syndrome a low CT-value, below 33 − 34 is expected. The inverse statement, that given a low CT-value of below 33 a patient is probably in the recovery phase, and not likely to be infectious, is by no means true. Thus, the need to prevent the spread of SARS-CoV-2 requires the use of precise detection methods, that are sensitive to viral loads of the order of 10 viral RNA units in a given sample. We discuss now how this issue can be addressed in the pooling test method.

To quantify the sensitivity variation that is caused by sample dilution we analyze the distribution of CT-values from tests collected in Dezember 2020 in the “Diagnostic laboratory for clinical pathology” in Hamburg. We chose a representative sample that contains samples from patients showing acute symptoms and positive tests obtained from tests of asymptomatic individuals.

Figure 3 shows the collected data from 795 patients in a Histogram, the applied detection technique is the automated RNA extraction procedure, and RT-PCR diagnostic provided by the *cobas*^®^ 6800 system developed by Roche^®^. The blue points indicate the predictions from our best-fit model to the data, and the magenta curve shows the underlying probability density function. The fitted model is a bi-normal distribution with a probability density given by *N*_Bi_ = *N* (*µ*_1_, *σ*_1_) *p* + (1 − *p*)*N* (*µ*_2_, *σ*_2_), with *N* (*µ, σ*) being the normal distribution. The best fit parameters are *p* = 0.65, *µ*_1_ = 23.3, *µ*_1_ = 31.9, and *σ*_1_ ≈ *σ*_2_ ≈ 3.7. The overall goodness of fit is given by a *χ*^2^ per degree of freedom of 1.2, which indicates an excellent description of the data set.

**FIG. 3.**
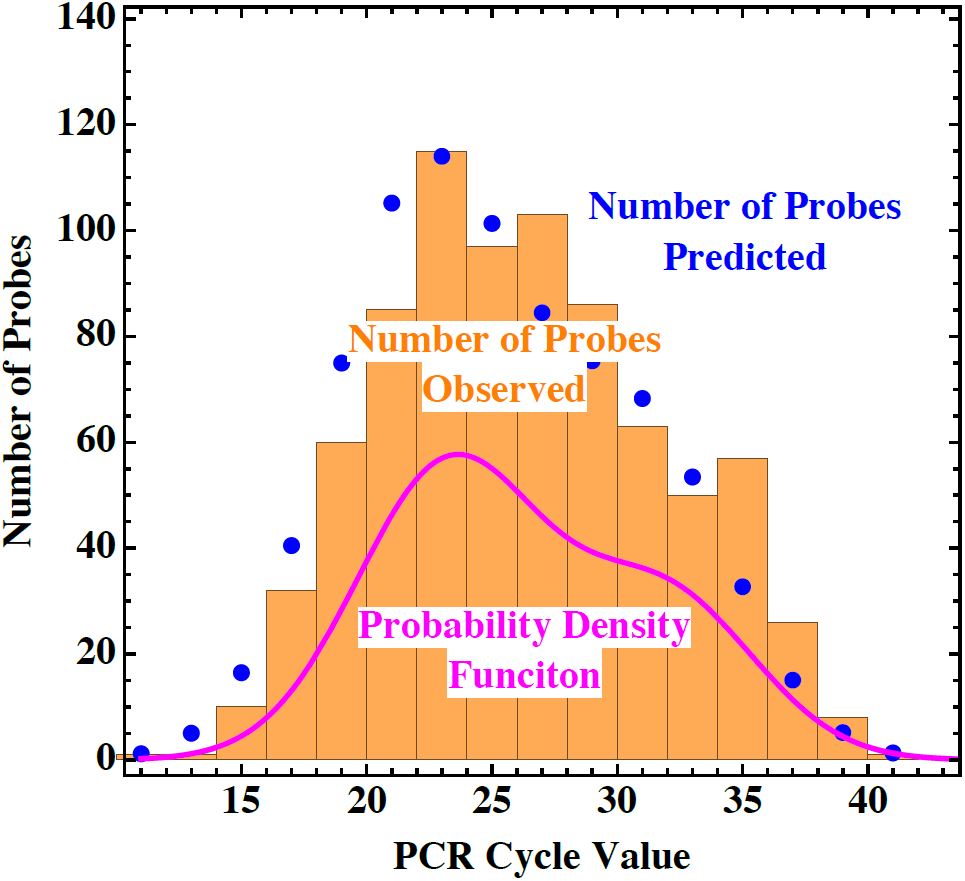
Panel b) shows the same data set as a histogram. The magenta line shows the theoretically fitted probability distribution and the blue dots show the integral values of the theoretical distribution for each bin.

The rate of false positive results is found by integrating the probability density function *N*_Bi_ over the range of CT-values above a defined detection threshold *T*, and is given by 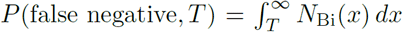. As an example, given the observed CT values distribution, we estimate the expected sensitivity of the Roche^®^ rapid Antigen test, which effectively detects samples corresponding to a *CT* ≈ 27 [12, 13]. The integral of the CT-value probability distribution yields a false negative probability of *P*_rapid_(false negative, 27) ≈ 40%, that is substantially larger than the systematic uncertainty of sample collection.

The detection threshold of the RT-PCR used in our study is at a CT value of 38. The expected rate of falsenegative probes is then given by *P* (false negative, 38) ≈ 2%, much smaller than the systematic uncertainty of about 10% estimated above. Given a dilution factor of a probe *D*, the false negative probability is *P* (false negative, 38 − log_2_(*D*)).

In Table I the expected probabilities for a false negative result are summarized. The crucial observation is that exceeding a dilution factor of *D* = 10 leads to a sensitivity loss that dominates the expected systematic sample-taking uncertainty. However, some strategies can be used to overcome this problem.

**TABLE I.**
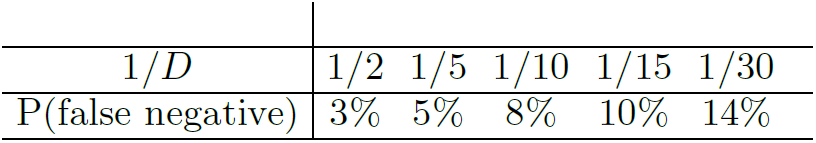
The dependence of the probability of a false negative result in a RT-PCR SARS-CoV-2 test given a dilution factor *D*.

| $1/D$ | 1/2 | 1/5 | 1/10 | 1/15 | 1/30 |
| --- | --- | --- | --- | --- | --- |
| P(false negative) | 3% | 5% | 8% | 10% | 14% |

- One method, used for the sensitivity improvement of pooled tests, was developed in [14]. The described procedure allows using RNA extraction to ensure the same viral RNA concentrations in the pooled test, as the would-be concentration in an individual test.
- A viable option for moderate pool sizes is extraction in a reduced buffer volume.
- Finally, a simple solution is the use of two test swabs, that need to be collected. One swab is used in the pool test and the second is kept for a possible after the test. Since the swabs are taken at the same time the viral shedding intensity in the patient is the same. The systematic uncertainty of a false negative result is in this case the same as in the individual testing method.

From a practical point, the above solutions have the following advantages and drawbacks. The first method is, clean, and allows to handle large patient pools. On the other hand, it requires substantial additional laboratory work. The second method is only applicable to moderate pool sizes with up to five test samples. The last method is simple and reliable, and only requires the collection of two test swabs from each patient.

In the experimental validation, we will thus make use of the second and third option, since they are most resource-conserving. An important question that remains to be answered is the optimal size of the test pool.

## Pool Size Optimization

The probability that at least one test in a given pool is positive and that repetition for all patients has to be performed, is given by

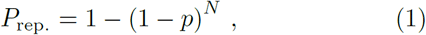

where *p* is the mean infection rate of the tested cohort and *N* the number of patients in a pool. If this possibility manifests itself additional *N* tests of the individual patients have to be performed. If the pool test is negative only one initial test is sufficient, such that the expected number of tests is given by *N*_total_ = (*N* + 1) *P*_rep._ + 1 − *P*_rep._ = 1 + *N P*_rep._. Given this construction, we can minimize the number of tests needed as a function of the pool size *N*, while keeping the infection rate *p* fixed. Our results, presented in the next section, allow us to optimize the pool size and adjust it to the patient cohort.

The function which determines the number of tests needed is numerically minimized with respect to the pool size. We find that the outcome strongly depends on the underlying infection probability.

In Fig. 5, we show the pool size optima for different assumptions for the mean infection rate of the cohort. At present, the mean infection rate, among the tests performed in Germany, seems to be around *p* ≈ 10% [15]. Thus a pooling of test samples without a cohort separation is not effective at the moment. However, we will demonstrate that an appropriate cohort selection can lead to a significant reduction in the testing efforts, and allows to dramatically increase the test capacities.

**FIG. 5.**
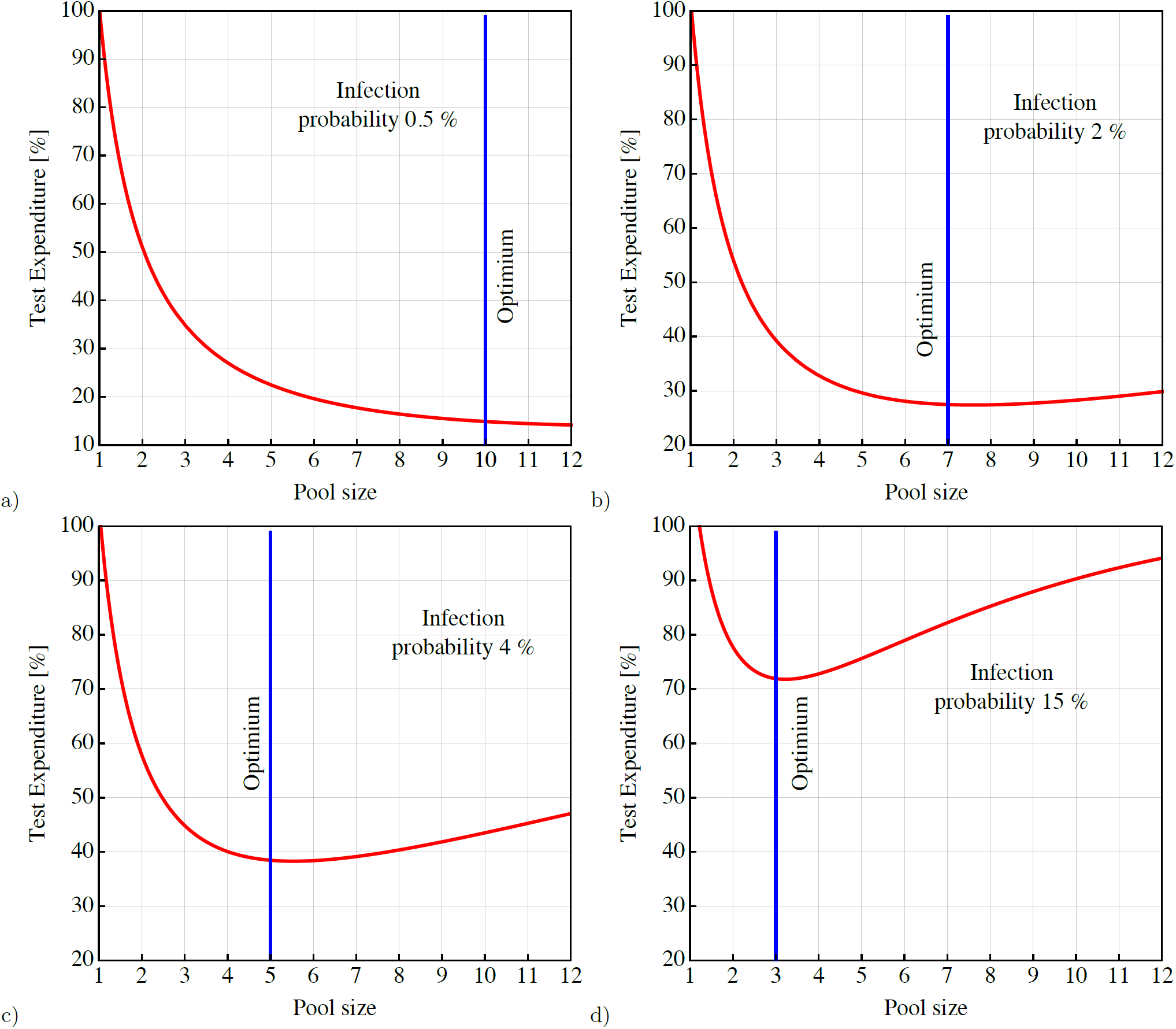
Optimal pool sizes, given a characteristic average infection rate a) *p* = 0.5%, b) *p* = 2%, c) *p* = 4%, d) *p* = 15%. In particular for low infection rates, a high efficiency improvement can be reached. Thus it can be highly beneficial for this method to identify cohorts with intrinsically low infection rates. Note that in the case of nearly degenerate minima it is advantageous to choose the smaller pool size, as this lowers the number of repeated tests, given the same total test number, and accelerates the average result times.

Table II shows the optimal pool sizes given a certain mean infection value for the tested cohort. It is shown that a good cohort selection with percent level infection rates can lead to an ∼ 90% reduction of the test effort. Thus, it is possible to increase the number of tested patients by an order of magnitude using the same number of tests.

**TABLE II.** The optimal pool sizes *N* given a certain mean infection number in a cohort *p*. For average infection rates above 10% the optimization becomes inefficient. However, for infection rates of 1% and lower the optimization factor exceeds 80%.

| $p$ | [%] | 0.1 | 0.5 | 1 | 2 | 3 | 4 | 5 | 6 | 7 | 8 | 9 |
| --- | --- | --- | --- | --- | --- | --- | --- | --- | --- | --- | --- | --- |
| $N$ | [Stk.] | 32 | 15 | 10 | 7 | 6 | 5 | 5 | 4 | 4 | 4 | 3 |
| Opt. | [%] | 94 | 86 | 80 | 73 | 67 | 62 | 57 | 53 | 50 | 47 | 44 |

## Experimental Validation

The laboratory validation of the method was performed in the period from May 5 2020 to December 15 2020. The test cohort were individuals from the region of Hamburg that showed no symptoms but was routinely tested to ensure safety in health care facilities, ship crews departing from the harbor of Hamburg, and companies screening employees working at the office.

For the pool size, five patients per pool were chosen. To reduce the dilution factor each throat swab was extracted in 2 ml Guanidinium thiocyanate buffer, then 400 *µ*l of the five samples were unified and tested by the *cobas* 6800 system. The standard extraction procedure of the *cobas* 6800 system uses 4 ml for the extraction of each swab. Thus the dilution of 1 : 5 was reduced by a factor of two and resulted in a total dilution of 1 : 2.5.

In the test period of 224 days, a total of 1910 pool tests were performed. The number of positive pool tests was 14 and thus resulted in 5 × 15 = 75 test repetitions to identify the individually positive patients. Therefore, a total of 1910 + 5 × 15 = 1985 tests were performed to test 9950 patients. This amounts to a reduction of the experimental effort by 79%. The percentage of positive tests in our test cohort was as low as *p*_tested_ = 0.15%. This was expected from the cohort choice and application in safety screenings. During the period of the study, the overall positivity rate of PCR tests across Germany was on average 3%, thus the risk factor of our cohort was twenty times lower than an average laboratory test in this period. Furthermore, we performed a likelihood analysis of the CT-values found in the pool tests, to estimate our dilution hypothesis.

Figure 4 shows the result of a Kolmogorov-Smirnov test of the pool test CT-values, that evaluated the probability that the numbers resulted from an underlying CT-value distribution determined in the previous section. The expected errors *σ*_1_ and *σ*_2_ were rescaled by the inverse square roots of the sample sizes, and the expectation values of the bi-normal distribution were shifted by a factor *µ*_*i*_ → *µ*_*i*_ + log_2_ *D* for *i* = {1, 2}. Thus, for example, a dilution by a factor *D* = 2.5 would result in CT-values that are larger by an additive factor of 1.3. The p-value curve shown in Fig. 4 has a maximum in the range of *D* = 3 − 4. Given the relatively small size of the positive pool sample, this is an excellent agreement with our initial hypothesis of a dilution factor of *D* = 2.5. We, therefore, can conclusively show the validity of the pooling procedure for the RT-PCR method for SARS-CoV-2, and given the bi-normal CT-value distribution model we find that its false-negative rate is below 5%, see Tab. I.

**FIG. 4.**
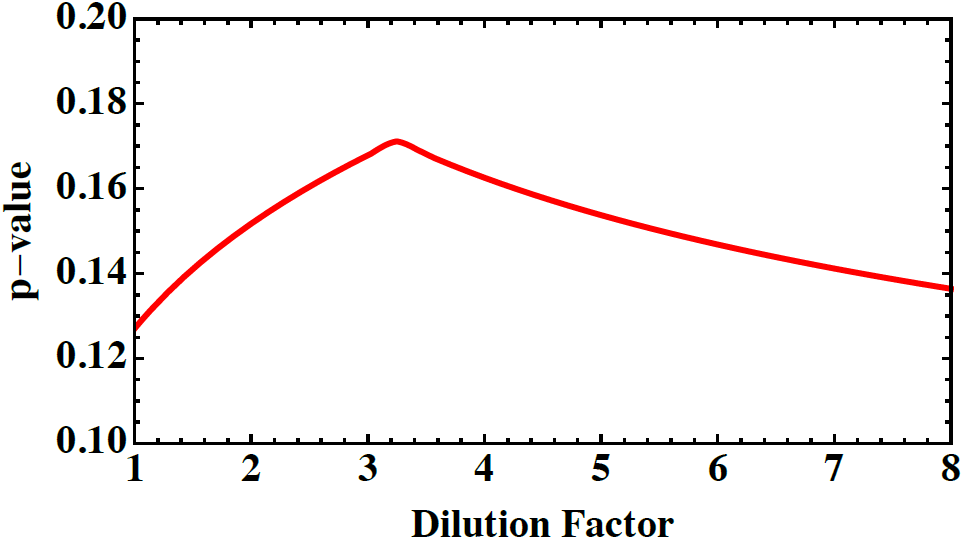
Probability values of a Kolmogorov-Smirnov test of the pool test CT-values. The maximum probability value indicates that the pool test CT-values are most likely to follow a Binormal distribution based on a shift of the dilution factor by a value of 3 to 4, which corresponds to a CT-value additive shift of 1.6 to 2.

## Risk Analysis

As discussed above the applicability and efficiency of the pool testing method are largely determined by the expected positivity rate of a given cohort. We adopt a simple hypothesis, that patients can be divided into risk groups. Those risk groups have intrinsic risk coefficients *r* that can be determined empirically and relate the positivity risk within a cohort to the total positivity rate of the population. We have thus a simple relation *R*_cohort_ = *R*_population_ × *r*.

We estimate the intrinsic risk coefficients based on our pool test study normalized to the average population positivity rate of the collection period in Hamburg, which was *R*_population_ ≈ 3% [16]. Furthermore, we use data collected from patients in the test center at the Hamburg main station ZOB for the weeks 44, 47, 48, 50 of the year 2020. The average population positivity rate within this collection period was higher at a value of *R*_population_ ≈ 9% [16].

Table III shows the estimated risk coefficients for five patient cohorts. Persons returning from trips within Germany considered as risk zones (Travelers), persons who received a warning by the COVID Warning APP (App-Warning), persons who had contact with an infected individual (Contact), persons who showed symptoms of Covin-19 (Symptom), and the cohort used for the pool test validation, tested in routine safety screenings (Screen).

**TABLE III.** Absolute numbers of tested patients, the positivity rate and the corresponding risk coefficients in five pre-defined patient cohorts.

|  | Travelers | App-Warning | Contact | Symptom | Screen |
| --- | --- | --- | --- | --- | --- |
| Total | 5428 | 1727 | 761 | 3046 | 9550 |
| Pos. | 184 | 30 | 43 | 516 | 14 |
| Rate | 3.4% | 1.7% | 5.7% | 17% | 0.15% |
| r | 0.4 | 0.2 | 0.5 | 1.9 | 0.05 |

Based on the determined risk coefficients and the current overall positivity rate of the population in Hamburg *R*_population_ ∼ 10% following pool sizes appear optimal.

- Returning travelers, and persons with a suspected contact with an other positive individual: Risk: *R*_population_ × *r* ≈ 4 %, Optimal pool size: *N* = 5.
- Corona App warning: Risk: *R*_population_ × *r* ≈ 2 %, Optimal pool size: *N* = 7.
- Routinely screened individuals: Risk: *R*_population_ × *r* ≈ 0.5 %, Optimal pool size: *N* = 15.

Individuals with symptoms resembling Covid-19 syndrome should not be tested in a test sample pool. This estimates are an example of how given a population positivity value *P*_population_, the risk coefficients in Tab. III can be used to identify the ideal test pools size using Tab. II. Finally note that to avoid sensitivity loss in pool tests with more than five patients, the double swab technique is highly recommended and represents the most simple solution to the sensitivity loss problem. We have successfully applied the double swab collection technique for safety screening of the personnel employed by the”Diagnostic laboratory for clinical pathology” in Hamburg.

## Summary

The current situation of the SARS-CoV-2 pandemic is highly worrisome. A potent weapon in the struggle for containment are systematically performed molecular biological SARS-CoV-2 tests. In this way, infected individuals can be identified, quarantined, and treated as fast as possible. In this letter, we suggest a pooling method, with the help of which the test capacities can be significantly increased. We demonstrate optimal pool sizes based on the underlying individual mean infection numbers. Also, we validate our theoretical predictions in a defined cohort of pooled patient tests.

We observe that the pool test efficiency strongly depends on the underlying rate of positive patients. We demonstrate a simple method of how to estimate the expected positivity rate of a given cohort based on the underlying population risk in a region. In our work, we use collected patient data to estimate the risk coefficients for five relevant patient cohorts. Ultimately, we note that the most promising application of the pool testing method is large safety screening in companies, medical facilities, and other critical institutions. Larger pool sizes will allow increasing the test capacity by more than 90%. In particular, since the sensitivity loss problem can be simply overcome by collecting two test swabs. One for the pool test, and one for a potential test to determine which patients were positive in the pool. With the method suggested here not only the test efficiency can be significantly improved, but also the cost of testing lowered and, time and valuable material resources preserved.

## Data Availability

External data for cycle values have been used from this source: He, X., Lau, E.H.Y., Wu, P. et al. Temporal dynamics in viral shedding and transmissibility of COVID 19. Nat Med 26, 672 675 (2020). https://doi.org/10.1038/s41591-020-0869-5

## Notes

### Competing Interest Statement

The authors have declared no competing interest.

### Funding Statement

The work of J.S. is largely Funded by the Alexander von Humboldt foundation.

### Author Declarations

The data of the study only contain PCR cycle values obtained in the routine diagnostic in the Institute for Clinical Pathology Dr. Fenner and Colleagues, which is accredited by the German Norm DIN EN ISO 15189.

